# Estimating the levels and trends of family planning indicators in 436 sub-national areas across 26 countries in sub-Saharan Africa

**DOI:** 10.1101/2021.03.03.21252829

**Authors:** Joshua L. Proctor, Laina D. Mercer

## Abstract

**Background:** Scaling up access to safe, effective, and voluntary family planning (FP) services to achieve universal access for women and families will require increased commitment by countries and international organizations. On the way toward universal access, quantitative family planning goals have also been established by the United Nations through the sustainable development goals (SDGs). Here, we present a model-based framework that can help monitor progress toward these goals at the sub-national and demographic subgroup scale.

**Methods:** We utilize 90 demographic health surveys for 26 countries that contain associated geographic position system data. We extract survey cluster level data to fit multiple small area estimation models that estimate FP indicators by administrative unit one and two regions as well as different demographic subgroups.

**Findings:** We find significant variation of modern contraceptive prevalence rates (mCPR), unmet need, and demand satisfied by country, sub-national region, and demographic subgroup. Our model-based estimates show that on average for 436 administrative unit one regions, mCPR has increased at a rate of 0.75% per year and unmet need has decreased by 0.26% per year. There are also striking differences of FP indicators by demographic subgroup; for example, unmet need can be up to 40% different based on age and parity.

**Interpretation:** We have developed a framework to help monitor progress, provide insights about the inequitable progress by region and demographic groups, and account for the sparsity of available data. These results and framework can help policy-makers better allocate and target interventions to help achieve family planning goals.

**Funding:** This publication is based on models and data analysis performed by the Institute for Disease Modeling at the Bill & Melinda Gates Foundation.

## 1. Introduction

Providing universal access to safe, effective, and voluntary family planning services is fundamental to ensuring the health and well-being of women and families around the world. Ambitious goals have been established in the last decade to meet the current demand for family planning services. These include goals such as Family Planning 2020 (FP2020), which aimed to increase the number of modern contraceptive users by 120 million by the year 2020 [1]. FP2020 was adopted by a wide coalition of governmental and non-governmental agencies in 2012; the international commitment by governments and non-governmental to FP2020 have translated to encouraging progress [2, 3]. In addition, 193 Member States of the United Nations agreed to the 2030 Agenda for Sustainable Development. The Agenda contains 17 Sustainable Development Goals (SDGs) [4] including a proposed target to achieve at least 75% demand satisfied for family planning with modern contracep-tives by 2030 [5]. Establishing quantitative targets for ambitious family planning is essential for galvanizing governmental action and focusing policy, advocacy, and intervention efforts within countries.

Setting and meeting ambitious quantitative targets for family planning requires a comprehensive monitoring and evaluation plan for key family planning indicators. As the *120 by 2020* goal was being developed, the cadence of nationally representative surveys, such as the Demographic Health Surveys (DHS), was identified as a limitation to monitoring the expected change of family planning indicators. A new survey instrument called the Performance, Monitoring, and Accountability (PMA) survey [6], was created to measure indicators at shorter intervals [1]. In addition, statistical models were developed that integrate data from multiple survey instruments, including PMA and DHS [7], to produce estimates at the national level for family planning indicators [8]. The family planning estimation tool (FPET) has been an integral innovation for tracking progress to-ward global goals [9]. Nonetheless, the recent model-based estimates indicate that the international coalition has fallen short of achieving *120 by 20* [3]; similarly, progress toward meeting SDG 3.7.1 by 2030 is not on track for success [10].

Despite the lack of progress, a sustained international commitment and a focus on family planning interventions can help accelerate progress toward family planning goals [11]. Well-established family planning interventions such as information, education, and communication (IEC) campaigns and post-partum family planning have demonstrated repeated success across multiple settings [12, 13]. However, many family planning interventions are typically implemented at sub-national geographic scale or targeted at specific demographic groups [12, 14]. In order to monitor and evaluate changes at these granular geographic and demographic resolutions, we have developed a novel, model-based framework that leverages data from nationally representative cross-sectional surveys to construct sub-national estimates for family planning indicators. Moreover, we demonstrate the statistical framework can estimate family planning indicators by different demographic sub-groups mirroring the scale and target of intervention efforts. This framework can be utilized by policymakers as a tool for retrospective and prospective planning.

Model-based frameworks have proven to be an important tool for monitoring family planning indicators. The FPET has been widely utilized to track progress for FP2020 countries [9] and is based on the foundational statistical modeling by Alkema et al. [8]. The model assumes a parametric function form of a logistic curve, colloquially referred to as an *s-curve*, which is based on the theory of social diffusion of ideas [15]. The scurve functional form also enables the statistical model to make long-term forecasts of indicators beyond recent available survey data. However, the strong underlying assumption of a logistic functional form can influence the estimated levels and trends of family planning indicators and is not consistent with data across all settings, especially where progress has stalled or declined. An alternate modeling approach, leveraging small area estimation techniques and complex survey design, has also been used to estimate family planning indicators in Nigeria [16]. This Bayesian methodology can help estimate the levels and trends of health care indicators at a subnational scale by leveraging solely survey data without imposing a functional form [17, 18]. In this work, we use this approach to estimate and forecast the levels and trends of family planning indicators across 26 countries in sub-Saharan Africa; we also highlight the differences in these indicators across demographic subgroups such as parity, age, and urban-rural. We also provide estimates of the change of mCPR and demand satisfied from 2010-2020.

## 2. Data and methods

### 2.1. Cross-sectional survey data

Survey data comes from 90 DHS for 26 countries. The DHS are nationally representative household survey with a multistage cluster design. Here, we utilize the DHS that contain associated geographic position system (GPS) data; this enables us to accurately associate clusters according to current administrative boundaries within each country. Supplement §1. provides specific details about which countries, surveys, complex design, and any identified inconsistencies across surveys. Figure 1.A.i. illustrates the 26 countries that are analyzed including how many surveys are associated with country; note that we only consider countries with two or more surveys, associated GPS data, and access to the individual level data.

**Figure 1.**
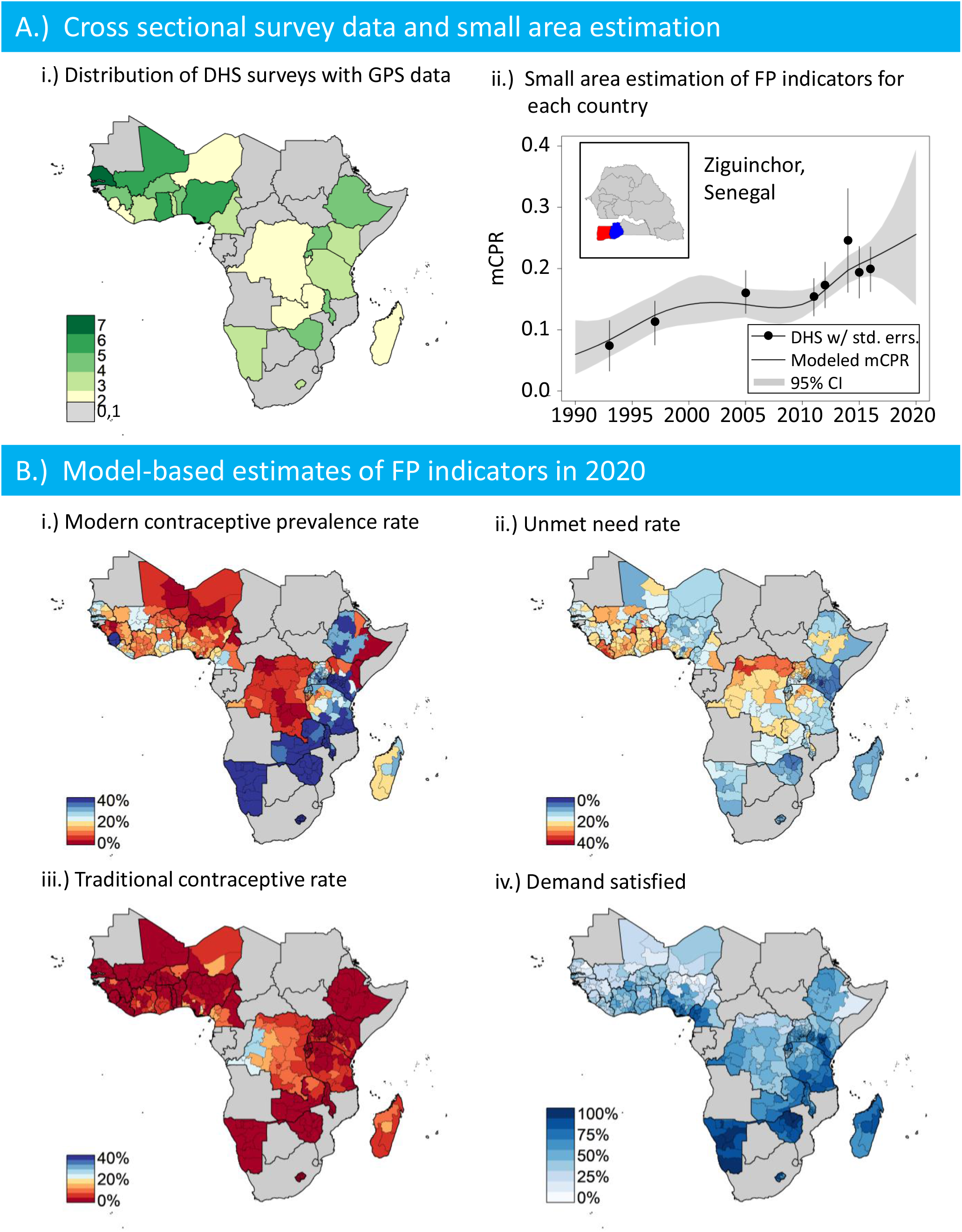
A.i) An illustration of showing how many DHS surveys with GPS data are available for each country. A.ii) Direct survey and model-based estimates with uncertainty intervals for Ziguinchor, Senegal from 1990-2020. B.) Model-based mean estimates of FP indicators in 2020 for B.i) modern contraceptive prevalence rate, B.ii) unmet need, B.iii) traditional contraceptive rates, and B.iv) demand satisfied.

For each survey, we extract individual level data describing modern contraceptive usage, traditional contraceptive usage, and unmet need. The revised definition of unmet need was used for all surveys [19]. We also extract demographic information such as age, parity, urban-rural residence, survey design variables, and GPS location information. Global administrative areas (GADM) shapefiles version 3.6 for administrative boundaries one and two are used in this analysis [20]. The GADM shapefiles enable the small area estimation analysis and illustrations of analytic results as sub-national maps of 26 countries. The github repository [21] contains scripts in the computing language R to perform the data extraction, analysis, and visualization of results.

### 2.2. Estimating rates of contraceptive usage, unmet need, and traditional contraceptive usage

Modern and traditional contraceptive prevalence rates were estimated by computing the proportion of women or partners using at least one method. Unmet need was computed by calculating the proportion of women that did not want to become pregnant, for the purpose of spacing children out or limiting all together; note that we are using the revised definition of unmet need for all surveys analyzed in this work [19]. Each of these indicators were estimated at the administrative unit level 1 and 2 across all 26 countries. In addition, we analyze these rates for different demographic subgroups of women. We analyze the rates for all women as well as partitioning women by eight different subgroups: nulliparous women age 15− 24, parous women age 15− 24, nulliparous women 25 years or older, parous women 25 years or older, urban women 15− 24, rural women 15− 24, urban women 25 years or older, and rural women 25 years or older. The direct estimates and design-based variances were calculated using the R survey package [22]. As an example, Figure 1B. shows the direct estimates of mCPR values and 95% confidence intervals for Ziguinchor, Senegal.

### 2.3. Models for sub-national estimation of family planning indicators

In this work, the spatio-temporal statistical models assume there is an underlying rate of FP indicators and treats the direct survey estimates as measurements with uncertainty. The models are fundamentally Bayesian and hierarchical allowing for multiple surveys, the inclusion of the complex survey design, and the explicit incorporation of survey estimate uncertainty; this model framework is similar to that found in [16]. In the first of two stages, the pseduo-likelihood is defined by the asymptotic distribution of the log transformed design-based estimates and transformed variance. At the second stage, the logit transformation of the assumed underlying true indicator are modeled linearly with independent random effects, random walks of order 1 and 2, and temporally structured space-time interactions. We also include survey-year and survey-geography random effects to account for systemic survey biases. In addition, independent spatially structured random effects are also included to provide geographical smoothing at the sub-national area resolution; the data and trends from physically adjacent areas are used to inform the spatial random effects. We construct multiple models with each of these components for each demographic subgroup. Supp. §3 describes each of the models. In this article, we report the medians and 95% confidence intervals from the posterior distribution. Figure 1B. illustrates the location of Ziguinchor, Senegal (red area in the inset chart), the area that is included in the spatial smoothing term (blue area in the inset chart), and the model-based median estimates and the confidence intervals.

### 2.4. Fitting the models to data

The Bayesian spatio-temporal model is fit with the integrated nested Laplace approximation (INLA) [23] within the R scientific computing language [24] using the INLA package [25]. The INLA has been shown to be an accurate and computationally efficient alternative to using Markov Chain Monte Carlo [26, 27]. We independently fit each of the Bayesian models with the same hyper-parameter priors. All R scripts for the analyses are available at the github repository [21].

### 2.5. Model selection procedure to evaluate

For each country and demographic subgroup, we use a principled model selection procedure that balances *goodness-of-fit* and *parsimony*. Supp. §3 outlines the various model components and structure. We implement three separate model selection procedures including the log conditional predictive ordinate (LCPO) [28], Watanabe-Akaike information criterion (WAIC) [29], and the deviance information criteria (DIC) [30]. Despite there being a lack of consensus on the best model selection criterion for spatio-temporal models [18], we utilize all three to evaluate the best model for each country and demographic subgroup. If there are inconsistencies across the selection procedures, they are resolved on a case by case basis through the direct inspection of model fit and forecast trend.

## 3. Results

### 3.1. A Bayesian hierarchical model reveals sub-national heterogeneity by geography and year

Model-based estimates for family planning indicators shows significant heterogeneity across and within 26 countries in sub-Saharan Africa. Sub-national estimates within western African countries generally have less than 20% mCPR, whereas eastern African countries have rates above 40% with countries such as Namibia and Kenya having greater than 50 (Figure 1B.i. and Supp. Figure 1). Northern Nigerian states are uniformly below 10% mCPR in comparison to southern states which are up to 20%; Senegal and Tanzania both have within country variability greater than 15% (Supp. Figure 1.). Unmet need does not vary across the 26 countries as much as mCPR ((Figure 1B.ii. and Supp. Figure 1)) However, estimates of within country variation of unmet need for 2020 is still approximately 10% for countries such as DRC, Uganda, Namibia, Lesotho, an Nigeria (Supp. Figure 1). Estimates of traditional contraceptive rates are uniformly low across all of the countries with very little variation within country with the exception of the DRC (Figure 1B.iii.).

Quantitative and qualitative estimates of demand satisfied varies significantly across sub-Saharan African countries (Figure 1B.iv.). Similar between and within country trends observed for mCPR and unmet need are reflected in demand satisfied. For example, the north and south of Nigeria have significantly different rates of demand satisfied (Figure 1B.iv.). Eighty-six sub-national areas have demand satisfied over 75%, whereas 226 areas have less than 50%. Moreover, 66 areas have less than 25% demand satisfied (Figure 1B.iv.).

### 3.2. Sub-national, country-specific models demonstrate cross border consistent estimates

Despite the sub-national models being country specific, the model-based estimates are consistent across borders. For example, Figure 1B.i. visually demonstrates low mCPR values across the eastern areas of Somalia and Kenya as well as the northern and southern areas of Nigeria and Niger. Similarly, Zambia, Malawi, and south eastern Tanzania all have higher values of mCPR. Values of unmet need are also consistent across western Africa countries such as Liberia, Cote d’Ivoire, Ghana, Togo, and Benin (Figure 1B.ii.). There is also a consistent unmet need values across Rwanda, Burundi, north western Tanzania, and an eastern area in the DRC.

### 3.3. Model-based estimates enable the assessment of the change in family planning indicators including demand satisfied from 2010-2020

Sub-national models of family planning indicators enable the estimation of the change in indicators from 2010 to 2020. Figure 2 shows the distribution of percent change per year for mCPR, unmet need, traditional contraceptive rates, and demand satisfied for 436 areas. The mean values are 0.75% change per year for MCPR, −0.26% change for unmet need, and 0% change for traditional contraceptive rates. There are 38 areas in sub-Saharan Africa that showed a greater than 2% change per year. Supp. Figure 1 shows how each countries mCPR and unmet need values change from 2010 to 2020. The distribution for unmet need is more centered around 0% with a substantial number of areas both increasing and decreasing unmet need over the decade (Figure 2).

**Figure 2.**
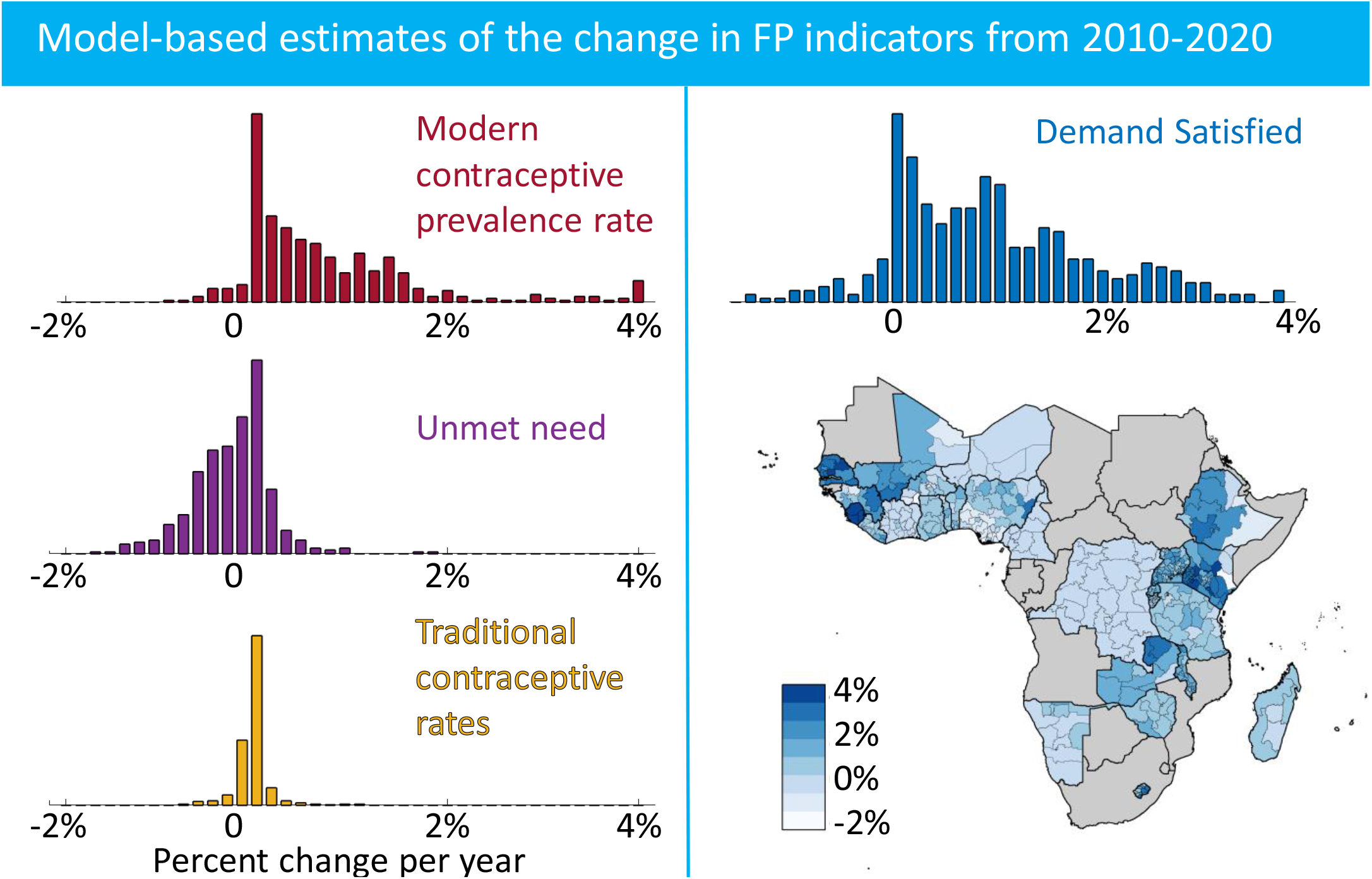
Model-based estimates of the percent change per year for family planning indicators over the 2010-2020.

Demand satisfied from 2010 to 2020 has broadly increased across much of sub-Saharan Africa. There are 184 areas with greater than 2% change per year of demand satisfied. Figure 2 illustrates the geographic distribution of change in demand satisfied. Senegal, Kenya, Uganda, and Sierra Leone have each had a significant increase in demand satisfied. Notably, there are 98 geographic areas with less than 0.1% change per year distributed across Nigeria, Guinea, Burkina Faso, Kenya, and Ethiopia.

### 3.4. Substantial heterogeneity exists in family planning indicators when partitioning women into demographic subgroups

Family planning indicators have substantial heterogeneity both sub-nationally as well as by demographic subgroup. The model-based framework enables estimation of indicators by groups such as different parity, age, and urban-rural.

#### 3.4.1. Parity and age subgroups

Parity and age are two key demographic characteristics that describe the heterogeneity in rates of contraceptive usage and unmet need among women. The difference in unmet need across the 26 countries analyzed in the article is quantitatively and visually striking; see Figure 3B. for a comparison between unmet need estimates for women aged 15− 24 without children and with children. Unmet need is estimated to be greater than 35% for women 15 − 24 with children across several southern areas of Nigeria, Benin, Togo, Ghana, Cote d’Ivoire, and Liberia in the year 2020. Modern contraceptive usage also varies significantly by age and parity (Supp. Figure 2); for example, women aged 15-24 without children are using modern contraceptives at a much lower rate than women with children over the age of 25. The southern areas of Uganda and Kenya all have consistently high (greater than 40%) mCPR for women with children in comparison to women without children (Supp. Figure 2). A complete table of model-based estimates from 2000-2020 is provided for these parity age groups in Supp. §3.

**Figure 3.**
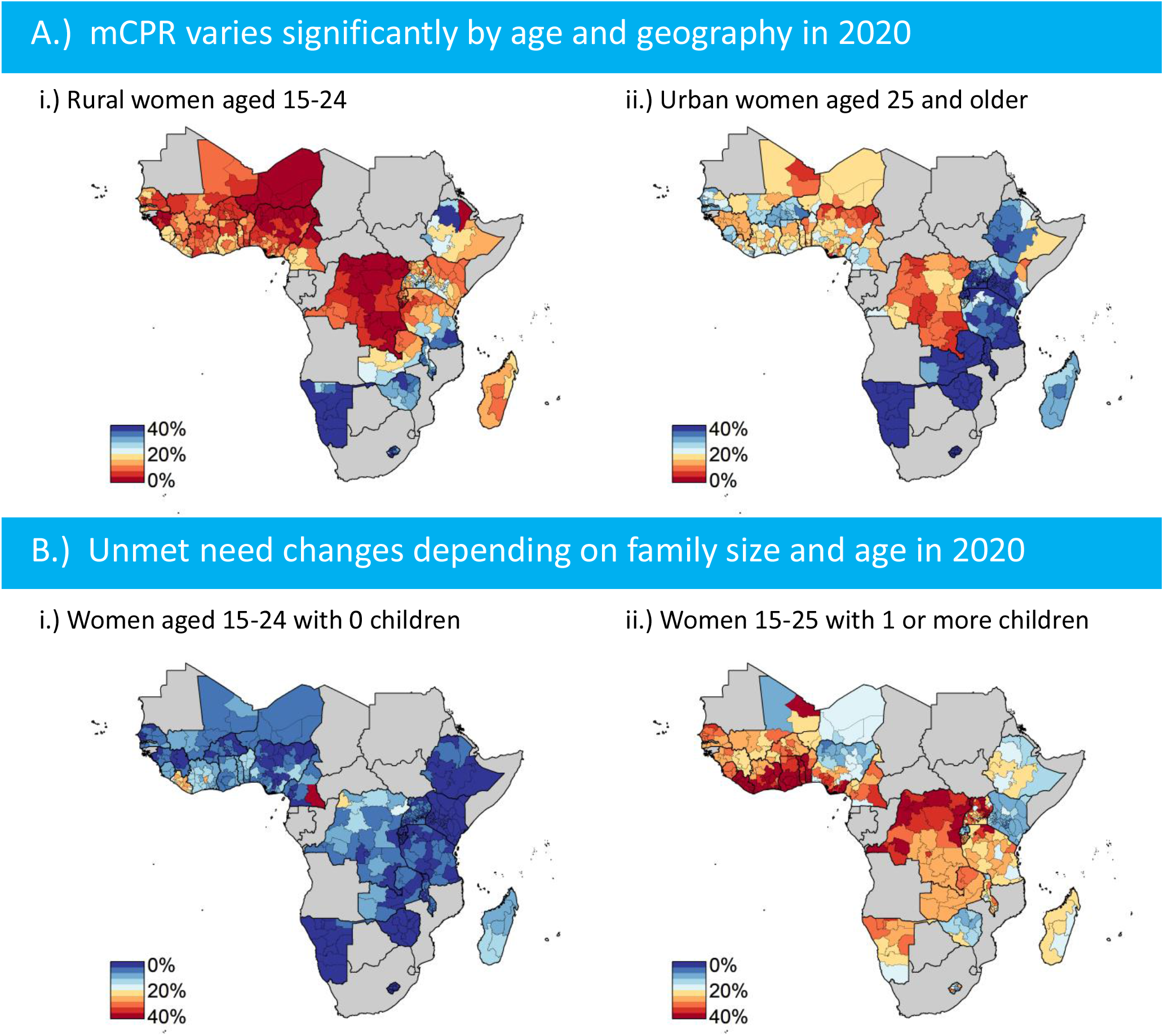
Model-based estimates of FP indicators in 2020 for modern contraceptive prevalence rates for women that are A.i.) rural and aged 15-24 and A.ii.) urban and aged 25 or older. B.) highlights estimates for unmet need for women that are B.i.) nulliparous and aged 15-24 and B.ii.) parous and aged 15-24.

#### 3.4.2. Age and urban-rural subgroups

Similar to parity, estimating family planning indicators by age and urban or rural residence reveals a high-level of heterogeneity. In general, Figure 3 illustrates that women in urban settings that are 25 or older tend to have much higher rates of contraceptive usage than younger women in rural areas. Subnational areas in several countries, including Nigeria, Uganda, Ethiopia, and Tanzania, show variability for each demographic subgroup (Figure 3). In western Africa, unmet need is higher for women aged older than 24; moreover, women living in rural settings are expressing more unmet need in countries (Supp. Figure 3).

### 3.5. Thies, Senegal: a case study in evaluating the levels and trends by demographic subgroups

Thíes, Senegal provides an important case study for understanding the magnitude of heterogeneity by sub-national area and demographic subgroups. Areas across Senegal have seen a substantial rise in mCPR and demand satisfied (Figure 1). Figure 4A. shows model-based estimates of mCPR from 1990 to 2020 for Thíes Senegal for all women; the clear increase in mCPR can be seen with the direct estimates from the continuous DHS as well as the model-based estimates. The levels and trends of mCPR by demographic subgroup vary by age and urban-rural residency. Women that are 15 − 24 and living in an urban setting have a low value of mCPR relative to all women (Figure 4B.)); in comparison, women older than 24 in rural settings have had a much higher adoption of modern methods from 2010-2020. Notably, the mCPR trend for younger women in rural settings shows a consistent uptick, which is not observed for urban women in the same geographic area. Similar time-series plots are available for all 436 sub-national areas by each demographic subgroups [21].

**Figure 4.**
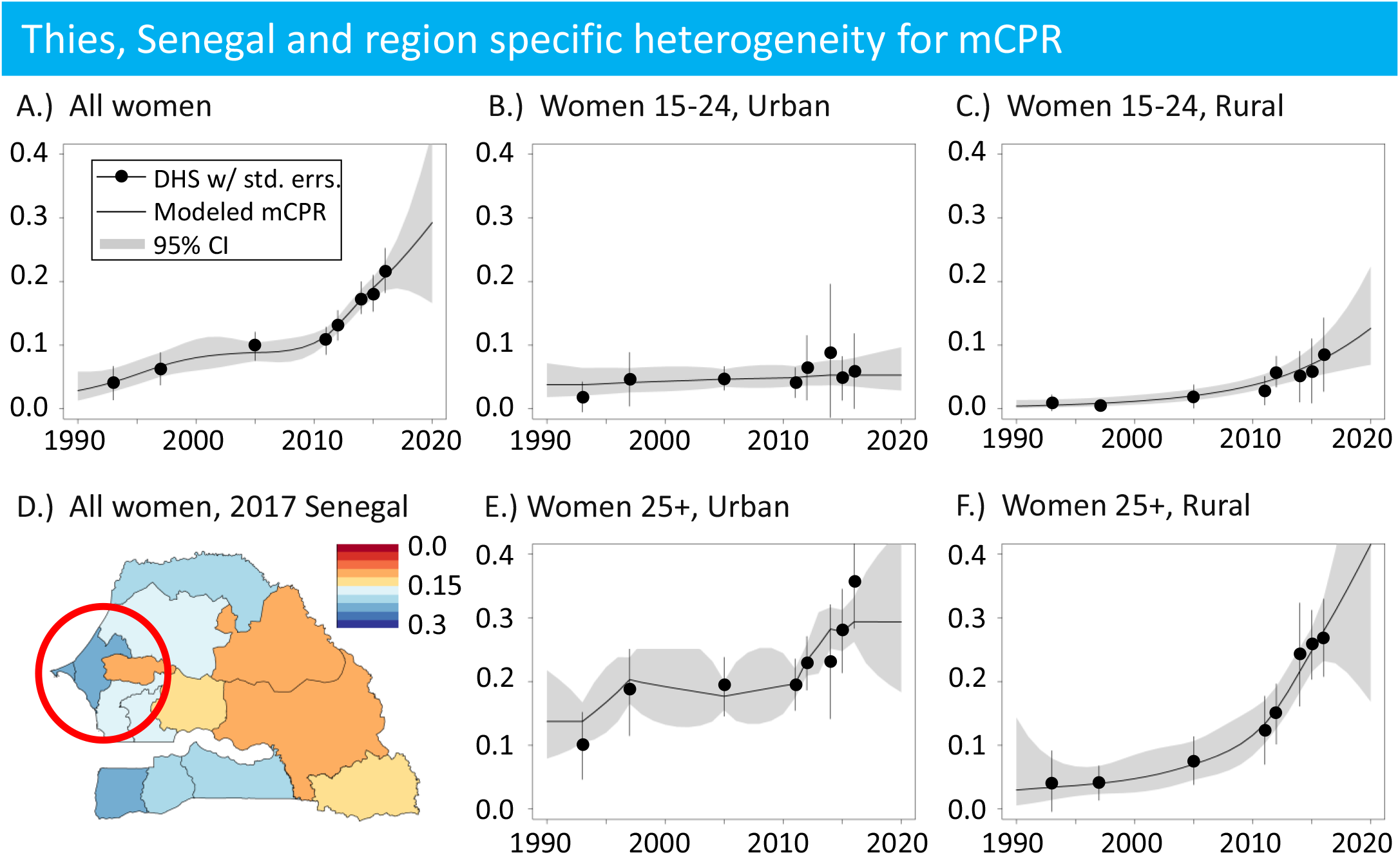
Direct survey and model-based estimates of mCPR in D.) Thìes, Senegal from 1990-2020 for A.) all women, B.) urban women aged 15 − 24, c.) rural women, aged 15 − 24, E.) urban women, 25 and older, and F.) rural women, 25 and older.

### 3.6. Model-based estimates of FP indicators at the administrative unit two regional level

Model-based mean estimates of FP indicators at the administrative unit two level are qualitatively similar to level one (Figure 5). For example, the estimated average yearly increase of mCPR and unmet need from 2010 to 2020 is 0.82 and − 0.22, respectively. The increased geographic resolution provides more insight into larger administrative unit regions such as Kaduna in Nigeria and northwestern Tanzania. However, the uncertainty intervals for each estimate is substantially wider (the mean estimates and 95% confidence intervals are reported in Supp. §4) with some administrative units having zero direct observations from survey instruments. This is especially true for countries with a large number of administrative 2 units and a small number of surveys like Nigeria.

**Figure 5.**
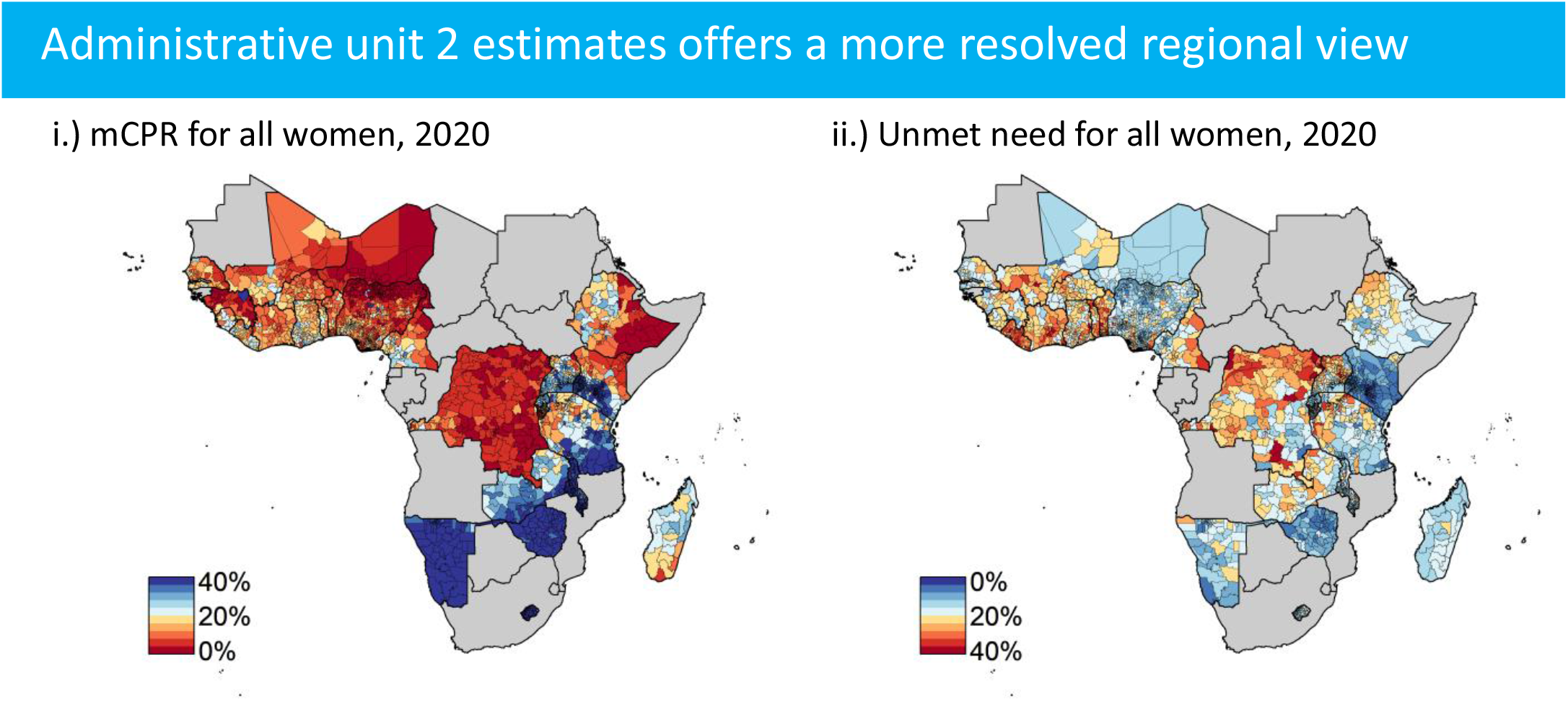
Model-based estimates of mCPR i.) and unmet need ii.) for all women in the year 2020 at the second administrative unit.

### 3.7. Model selection and sensitivity

We performed a series of model selection and sensitivity analyses to evaluate the robustness of the model fitting and results. For model selection, we identify the best-fit model, as outlined in §2.5; Supp. Table 5 provides the country-specific model chosen and the computed values of each selection metric. The model selection procedure was able to select the more complex random walk models for five countries; note that these countries tended to have more survey data available than others. The CPO criteria selected models that estimated unrealistic increases of mCPR after the last survey point for three countries. For each of these cases, we typically selected the random walk one models which typically had a similar model selection metric value; this helped constrain model-based predictions of mCPR increase outside of what has been previously observed in survey data. This heuristic procedure did not affect the model selection procedure for countries with clear increases in mCPR, such as Senegal depicted in Figure 4. The Bayesian framework also enables forecasts of family planning indicators with associated uncertainty quantification The uncertainty grows the further away in time from the last survey data point; see Figure4 for a clear illustration.

## 4. Discussion

We have constructed a modeling framework to generate estimates of modern contraceptive prevalence rates, unmet need, traditional contraceptive rates, and demand satisfied at a subnational geographic scale across 26 countries in sub-Saharan Africa. The framework leverages small area estimation techniques and Bayesian hierarchical modeling to quantify uncertainty for each estimate, incorporate survey sampling and non-sampling errors, and include spatio-temporal random effects. In addition, the framework enables quantification of the differences in rates of modern contraceptive prevalence rates and unmet need across demographic subgroups. We also generate short-term forecasts of indicators allowing for estimation of the change in FP indicators from 2010-2020. Combined, our approach provides public health officials a quantitative characterization of FP indicators at a subnational and demographic scale that could help inform the targeting of family planning programs.

Our results are broadly consistent with other model-based studies of FP indicators. Previous efforts have leveraged Bayesian hierarchical models to estimate FP indicators at the national scale [8] and is the basis of the widely used FPET [2]. FPET, though, is fundamentally lacking in it’s ability to estimate FP indicators sub-nationally. SAE techniques have also been previously used to estimate indicators within Nigeria utilizing multiple survey instruments [16]; in addition, SAE techniques have been applied to data from ten LMIC countries using the non-nationally representative Performance, Monitoring, and Accountability surveys [31]. Mercer et al. also demonstrated the methodology can be applied to demographic subgroups identified within the survey [16].

The results in this article also significantly broadens the scope of previous work. The model-based estimates of family planning indicators at the first and second administrative unit across 26 countries in sub-Saharan Africa helps quantify the differences across and within countries. We find substantial differences in contraceptive rates across urban-rural, age, and parity demographic subgroups with differences up to 30% in mCPR and 40% in unmet need. We have also quantified the change in FP indicators from 2010 to 2020. Demand Satisfied has overall increased across sub-national areas; however, several areas have also decreased over the last decade. This follows from unmet need increasing, potentially due to additional demand, driving demand satisfied lower. Our results help quantify this change over time and provide a basis for forecasting an expected change over the coming decade.

A demographically resolved characterization of FP indicators provides more insight into the levels and trends of FP indicators in a sub-national region. Here, we focused on a case study in Thíes, Senegal. The general increase in contraceptive prevalence at the national level and within Thie`s for all women is not equitably represented across demographic subgroups. We find that younger women from urban settings have a qualitatively different trend from their rural and older counterparts. This may seem counter to broader trends of contraceptive uptake occurring in urban settings before rural, but the results reflect the current stigma of contraceptive usage by younger urban women. The modeling framework has enabled this descriptive deep dive and is available for all geographic regions.

Our approach is limited by the amount and extent of cross-sectional survey data. The demographic health surveys occur on average every five years, with notable exceptions such as Senegal where there is a yearly rolling survey. The SAE framework is data-driven relying heavily on measurement data for model-based estimates; the low cadence of surveys limits the temporal and geographic resolution of the model-based framework. Moreover, the SAE framework requires cluster level data to have associated GPS locations in order to estimate indicators below the first administrative unit. The DHS provides this geographic data, but other surveys such as MICS or PMA lack this geospatial resolution hindering the ability to include those survey instruments in the SAE framework as Mercer et al. [16] did for Nigeria at the state level. In addition, without a strong dependence on a functional form similar to FPET, forecasting into the future depends on the recent survey estimates. To mitigate these challenges, the SAE framework provides a procedure for quantifying the uncertainty due to either the small number of surveys, small geographic region, or forward prediction. Here, we have focused predominantly on the first administrative unit estimates since we have direct survey measurements for each region, but we also provide estimate and uncertainty intervals at the second for areas that may be of interest to public health officials. The posterior distributions provide practitioners the ability to compare differences statistically as well as visually communicate the fidelity of the estimates to stakeholders at both the first and second administrative resolution.

Setting and making progress toward global goals such as the SDGs will require a continued international commitment focused on policy, advocacy, and intervention efforts. As new intervention programs are implemented, such as post-partum family planning and targeted information, communication, and education programs, assessing the impact will become essential to demonstrate their effectiveness and provide evidence for broader investment and adoption. The model-based framework in the article provides an important tool for monitoring progress at the same geographic and demographic resolution of interventions Also, our approach helps quantify the heterogeneity across demographic subgroups. Highlighting the inequities in terms of progress across regions and groups can help inform public health officials as they develop and implement family planning strategies for their population.

## Data Availability

All data used in this study is available publicly.

## 5. Authors’ Contributions

JLP contributed to the conceptual design of the study. JLP and LDM contributed to the analysis of the survey microdata. JLP and LDM completed hierarchical modeling. JLP contributed interpretation of analysis. JLP created the figures and wrote the article. All authors have read and approved the final manuscript.

